# Altered neurobehavioral reward response predicts psychotic-like experiences in youth exposed to cannabis prenatally

**DOI:** 10.1101/2024.08.23.24312453

**Authors:** Carolyn M. Amir, Dara G. Ghahremani, Sarah E. Chang, Ziva D. Cooper, Carrie E. Bearden

## Abstract

**Importance:** Rates of prenatal cannabis exposure (PCE) are rising with increasingly permissive legislation regarding cannabis use, which may be a risk factor for psychosis. Disrupted reward-related neural circuitry may underlie this relationship.

**Objective:** To elucidate neural mechanisms involved in the association between PCE and youth-onset psychotic-like experiences by probing correlates of reward anticipation, a neurobehavioral marker of endocannabinoid-mediated dopaminergic function.

**Design, setting, and participants:** This longitudinal, prospective study analyzed task-related functional neuroimaging data from baseline (n=11,368), 2-year follow-up (n=7,928), and 4-year follow-up (n=2,982) of the ongoing Adolescent Brain and Cognitive Development (ABCD) Study, which recruited children aged 9 to 10 years old at baseline from 22 sites across the United States.

**Results:** PCE (n=652 exposed youth) is longitudinally associated with psychotic-like experiences. Blunted neural response to reward anticipation is associated with psychotic-like experiences, with stronger effects observed in PCE youth (all |β| > 0.5; false discovery rate [FDR]–corrected *P* < .05). This hypoactivation at baseline predicts psychosis symptomatology in middle adolescence (4-year follow-up visit; β=-.004; FDR-corrected *P* < .05). Dampened behavioral reward sensitivity is associated with psychotic-like experiences across baseline, 2-year follow-up visit, and 4-year follow-up visit (|β| = .21; FDR-corrected *P* < .001). Psychotic-like experiences are positively associated with trait-level measures of reward motivation and impulsivity, with stronger effects for PCE youth (all |β| > 0.1; all FDR-corrected *P* < .05).

**Conclusions and Relevance:** Blunted activation in reward-related brain regions may serve as a biomarker for disrupted reward processing and increased psychosis risk during development. PCE may affect childhood behaviors and traits related to altered reward sensitivity.

## 1. Introduction

Past-month cannabis use among pregnant people increased by over 130% from 2003 to 2022 in the United States with increasingly permissive legislation.^1,2^ While there is growing evidence for associations between early cannabis use and elevated psychosis risk, less is known about the effects of cannabis exposure *in utero*.^3–7^ Recently, prenatal cannabis exposure (PCE) has been linked to psychotic-like experiences in early childhood.^8,9^ While the neurobiology underlying the link between cannabis and psychosis is not fully understood, the brain’s reward pathways including the dopaminergic mesocorticolimbic pathway are implicated in both cannabis use disorder (CUD) and psychosis.^10,11^ It is unclear whether these pathways mechanistically link early cannabis exposure and psychosis-proneness.

Cross-sectional evidence in a large sample of youth (n = 11,489) suggests that PCE increases risk for psychotic-like experiences—subthreshold psychotic symptoms that precede and may predict the onset of overt psychotic disorder^12–14^—even in pre-adolescence.^8^ Resting-state functional magnetic resonance imaging studies (rsFC) studies have found that rsFC in the salience network mediates the association between PCE and child psychopathology in pre-adolescence,^15^ and PCE is longitudinally associated with striatal rsFC in children ages 9-12.^16^ However, functional neural correlates of the relationship between PCE and psychotic-like experiences have not yet been fully elucidated, nor have these relationships been modeled longitudinally. Further, behavioral and trait-level factors contributing to psychosis and substance use vulnerability including reward motivation and impulsivity have not yet been examined. Studying the relationship between PCE, psychotic-like experiences, and their reward-related neural and behavioral correlates is of particular clinical significance in adolescence, representing a crucial risk period for the onset of overt psychotic disorder and substance use vulnerability.^3,6,17^

We examine the relationship between neurobehavioral markers of reward anticipation in task-based functional magnetic resonance imaging (fMRI) from the Adolescent Brain and Cognitive Development (ABCD) Study, a population-based, longitudinal cohort of 11,878 children ages 9 to 10 at baseline. We analyze the reward anticipation phase of the Monetary Incentive Delay (MID) task, a task designed to probe reward response, as dopamine neurons in reward-related brain regions fire more readily to reward-predictive cues rather than reward delivery.^18^ This preferential firing during reward anticipation is thought to represent the motivational processing of incentive attributed to reward mediated by larger systems encompassing mesolimbic dopaminergic transmission.^19^ Further, hypoactivation during the anticipation phase of the MID task has been demonstrated in both adults with schizophrenia^19^ and adults with CUD.^20^

First, we aim to test whether the association between psychotic-like experiences and PCE persists into later adolescence. Next, the main hypotheses tested using baseline data (n=11,368) are that: 1a) psychotic-like experiences are associated with blunted neural activation in reward-related brain regions (i.e., striatum and ventromedial prefrontal cortex) in response to reward expectancy; and 1b) within these models, relationships are stronger for PCE youth. We then examine the prospective relationship between brain activation to reward-predictive cues at baseline and psychotic-like experiences at 4-year follow-up (middle adolescence, n=2,982). We hypothesize that: 2) blunted activation in reward-related brain regions in PCE youth at baseline in response to reward-predictive cues is associated with more severe psychotic-like experiences in middle adolescence. Third, we examine associations between psychotic-like experiences by PCE status in behavioral and trait-level measures of reward motivation and impulsivity across time points, hypothesizing that: 3a) behavioral response time to reward-predictive cues is associated with psychotic-like experiences, and differs by PCE status; and 3b) psychotic-like experiences are associated with greater trait-level reward responsiveness, drive, and impulsivity, with stronger effects for PCE youth.

## 2. Methods and Materials

### Data Source and Participants

Data came from children (n=11,368 at baseline; M±SD age = 9.83±0.6 years; 47.85% girls; 74.13% White), born between 2005 and 2009 in the ongoing longitudinal ABCD study (data release 5.1.0; https://abcdstudy.org/). All parents/caregivers and children provided written informed consent and verbal assent, respectively, to a research protocol approved by the institutional review board at each data collection site throughout the US (Supplementary Figure 1, Supplementary Methods, section 1, and https://abcdstudy.org/study-sites/). This included data from three study time points: baseline, 2-year study follow-up visit, and 4-year study follow-up visit.

### Outcomes and Measures

#### Psychotic-like experiences

Psychotic-like experiences were assessed using a summary score for level of distress from the Prodromal Questionnaire - Brief Child Version (PQ-BC)^21^, a 21-item, developmentally-appropriate questionnaire that was validated in the ABCD study sample (for more information, see Supplementary Methods, section 2).

#### Prenatal Cannabis Exposure (PCE)

PCE is based on parent or caregiver (9,709 of 10,716 biological mothers [90.60%]) retrospective report. Three mutually exclusive groups were formed (at baseline): no exposure (n = 10,716), exposure prior to maternal knowledge of pregnancy only (n = 420), and exposure after maternal knowledge of pregnancy (this group includes exposure before and after knowledge of pregnancy; n = 232. Supplementary Methods, section 3).

### fMRI Paradigm

ABCD imaging data collection, acquisition, and analysis have previously been described.^22–24^ The Monetary Incentive Delay (MID) Task is a well-validated probe designed to measure domains of reward processing, including anticipation and receipt of reward, and modulation of motivation.^25,26^ See Supplementary Methods, sections 1 and 4, and Supplementary Figure 2 for details on task design and data acquisition and analysis. We assessed neural activation in the striatum and ventromedial prefrontal cortex (vmPFC) during reward anticipation (Supplementary Methods, section 5).

### Statistical analyses

All models included age, biological sex, parental education, and income as covariates, with research site and family unit (nested within site to account for twins and triplets in this study sample) as random effects. To determine if differences in psychotic-like experiences between groups could be attributed to differences in other prenatal or environmental exposures,^27^ we included variables measuring birth weight, prenatal exposure to tobacco or alcohol (before or after maternal knowledge of pregnancy), unplanned pregnancy, and prenatal vitamin use in models testing group differences in the association between PCE and psychotic-like experiences. Imaging analyses additionally included mean framewise displacement as a covariate.^28^ Models testing the association between baseline measures and psychotic-like experiences at 4-year follow-up included baseline psychotic-like experiences as an additional covariate. Participant ID was included as a random effects term in longitudinal models. All results reported were false discovery rate (FDR) corrected for multiple comparisons. A q-value <0.05 was considered statistically significant. Results are expressed as standardized coefficients with 95% confidence intervals. See Supplementary Figure 1 and Supplementary Methods, section 5 for further details.

To address the question of whether associations between PCE and psychotic-like experiences persist through middle adolescence, we conducted linear mixed models with psychotic-like experiences as the dependent variable and cannabis exposure group (unexposed versus exposed, with unexposed as the reference group) as the independent variable longitudinally (see Supplementary methods, section 6 for detailed statistical methods).

Cross-sectional relationships between activation in reward-related brain regions during reward anticipation and psychotic-like experiences at baseline tested hypotheses that psychotic-like experiences (DV) are associated with (a) blunted activation in response to reward anticipation in *a priori* reward-related regions of interest (ROIs), and that (b) effects would be stronger for PCE youth. Linear mixed models test the associations between activation in *a priori* ROIs and psychotic-like experiences, with group-by-region interactions tested on each ROI (striatum and vmPFC), during reward anticipation (Supplementary Methods, section 1). As a control analysis, we re-ran these models with amygdala and insula ROIs; though not primary elements of this pathway, both regions are involved in the mechanisms of addiction, substance use, and reward-related decision-making. (Supplementary Methods, section 7).^29–32^

To test the hypothesis that hypoactivation in reward-related brain regions at baseline is prospectively associated with greater psychotic-like experiences in adolescence, linear mixed models with psychotic-like experiences at 4-year follow-up as the DV, and baseline activation in reward-related ROIs as IVs, were tested across groups. We utilize baseline neuroimaging data to establish the predictive validity of an early biomarker of elevated risk for developing psychotic-like experiences, potentially present as early as childhood, as a potential predictor of outcome at the latest available time point.

We analyze behavioral performance on the MID task (reaction time to reward-predictive cues) using longitudinal linear mixed effects models across all time points, with psychotic-like experiences as the DV and RT as the IV. We then test group-by-behavior interactions to test the hypothesis that the relationship between behavioral performance and psychotic-like experiences differs as a function of exposure status.

We then investigated the relationship between psychotic-like experiences and trait-level factors related to reward motivation and impulsivity by PCE status to test the hypothesis that psychotic-like experiences would be associated with these trait-level factors, with stronger effects for PCE youth. Longitudinal linear mixed models tested the associations between trait measures and psychotic-like experiences, with group-by-trait interactions tested on each trait measure. See Supplementary Methods, section 8 for detailed information on trait measures.

## 3. Results

### Participant Demographics

We analyzed task-based fMRI and behavioral data acquired during the Monetary Incentive Delay (MID) Task from participants in the Adolescent Brain and Cognitive Development Study (ABCD; https://abcdstudy.org) at baseline (n=11,368; Mage = 9.83±0.6, 47.85% girls), 2-year follow-up (n=7,479 unexposed youth; n=144 exposed after knowledge of pregnancy; n=305 exposed before knowledge of pregnancy), and 4-year follow-up (n=2,826 unexposed youth; n=61 exposed after knowledge of pregnancy; n=95 exposed before knowledge of pregnancy). Table 1 describes baseline demographics of our sample, stratified by PCE status. Supplementary Table 1 includes details on rates of participant substance use by study time point.

**Table 1:** Baseline Adolescent Brain and Cognitive Development (ABCD) Study Sample Characteristics. ^1^No Prenatal Exposure>Prenatal Exposure both Before and Post-parental knowledge of pregnancy ^2^No Prenatal Exposure<Prenatal Exposure both Before and Post parental knowledge of pregnancy ^3^Prenatal Exposure Post parental knowledge of pregnancy <both No Prenatal Exposure and Prenatal Exposure Before parental knowledge of pregnancy.

| Variable | Prenatal Cannabis Exposure Post Knowledge of Pregnancy (N=232) | Prenatal Cannabis Exposure Before Knowledge of Pregnancy (N=420) | No Prenatal Exposure (N=10,716) |
| --- | --- | --- | --- |
| Child Variables |  |  |  |
| Age in months at baseline, mean (SD) | 117.90 (7.44) | 119.05 (7.70) | 119.05 (7.49) |
| Sex, female, N (%) | 126 (54.3) | 199 ( 47.4) | 5123 (47.8) |
| Birth weight in kgs.,mean (SD) | 6.36 (1.53) | 6.63 (1.39) | 6.63 (1.39) |
| Race/Ethnicity, N (%) |  |  |  |
| White<sup>1</sup> | 144 (59.50) | 242 (58.60) | 8203 (75.72) |
| Black | 105 (43.39) | 170 (41.16) | 2089 (19.28) |
| Asian | 3 (1.24) | 3 (0.73) | 193 (1.78) |
| Pacific Islander | 0 (0.00) | 4 (.97) | 34 (0.31) |
| Native American | 14 (5.79) | 24 (5.81) | 351 (3.24) |
| Hispanic | 33 (14.2) | 90 (21.5) | 2191 (20.5) |
| Other | 9 (3.72) | 33 (7.99) | 733 (6.77) |
| Pregnancy and Family Variables |  |  |  |
| Unplanned Pregnancy, N (%)<sup>2</sup> | 164 (70.69) | 304 (72.38) | 3837 (35.81) |
| Prenatal Vitamin Use, N (%)<sup>3</sup> | 171 (73.7) | 379 (90.2) | 9872 (92.2) |
| Household Income, N (%)<sup>1</sup> |  |  |  |
| \$0-49,999k | 181 (78.02) | 307 (73.10) | 6267 (58.48) |
| \$50k-74,999k | 16 (6.90) | 50 (11.90) | 1577 (14.72) |
| \$75-\$99,999k | 11 (4.74) | 23 (5.48) | 859 (8.02) |
| \$100k-\$199,999k | 4 (1.72) | 12 (2.86) | 836 (7.80) |
| \$200k or more | 8 (3.45) | 3 (.71%) | 186 (1.74) |

### Psychotic-like Experiences by PCE Status

PCE was associated with more severe psychotic-like experiences longitudinally across three waves of data, relative to those who were unexposed (see Supplementary Table 2 for full results). Analyses assessing differences in youth exposed to cannabis before versus after parental knowledge of pregnancy revealed that cannabis exposure both before and after parental knowledge of pregnancy was associated with more severe psychotic-like experiences compared to the unexposed group, with no statistically significant differences between the two exposure groups (Supplementary Table 2).

### Relationships between Neural Activity During Reward Anticipation and Psychotic-Like Experiences at Baseline

Figure 1 shows the results of models testing the relationships between PCE, activation in *a priori* ROIs, and psychotic-like experiences in youth. As hypothesized, striatal activation during reward anticipation was inversely associated with reported psychotic-like experiences and was blunted in PCE youth compared to unexposed youth (Supplementary Table 3). While there was no significant association between vmPFC activation and psychotic-like experiences, vmPFC activation was blunted in PCE youth compared to unexposed youth. Results of secondary analyses revealed that psychotic-like experiences were not associated with amygdala or insula activation during reward anticipation.

**Figure 1:**
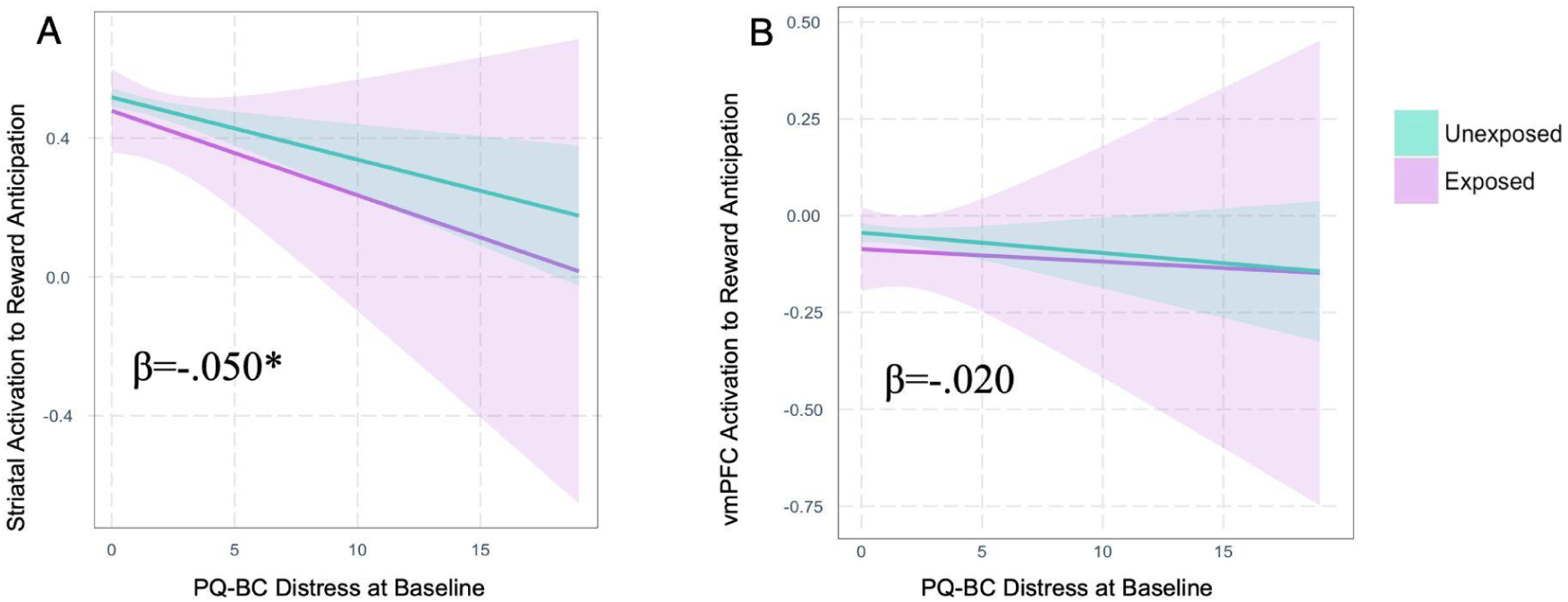
a. Baseline (cross-sectional) associations between psychotic-like experiences and activation in A) the striatum and B) the ventromedial prefrontal cortex during reward anticipation in the Monetary Incentive Delay Task by group status (exposed to cannabis prenatally versus unexposed). Striatal activation was inversely associated with reported psychotic-like experiences (A). There was no significant association between vmPFC activation and psychotic-like experiences regardless of PCE status (B). There were significant main effects of group; specifically, blunted activation in the striatum and vmPFC in youth exposed to cannabis prenatally during reward anticipation compared to unexposed youth. There were no significant group-by-region interactions. Shading around the best-fit lines indicates standard error. Covariates: age, biological sex, parental education, income, and mean framewise displacement as fixed effects, with research and family unit (nested within site to account for twins and triplets in this study sample) as random effects. Abbreviations: PQ-BC=Prodromal Questionnaire - Brief Child Version; vmPFC=ventromedial prefrontal cortex. Significance: *p<.05, FDR corrected.

### Association of Baseline ROI Activation with Psychotic-like Experiences at 4-Year Follow-up Visit

Figure 2 shows results of models using baseline fMRI data to predict psychotic-like experiences at 4-year follow-up. As hypothesized, activation in the striatum during reward anticipation at baseline was associated with greater psychotic-like experiences at 4-year follow-up across groups, despite controlling for baseline psychotic-like experiences. However, consistent with cross-sectional findings, baseline vmPFC activation was not significantly associated with psychotic-like experiences.

**Figure 2:**
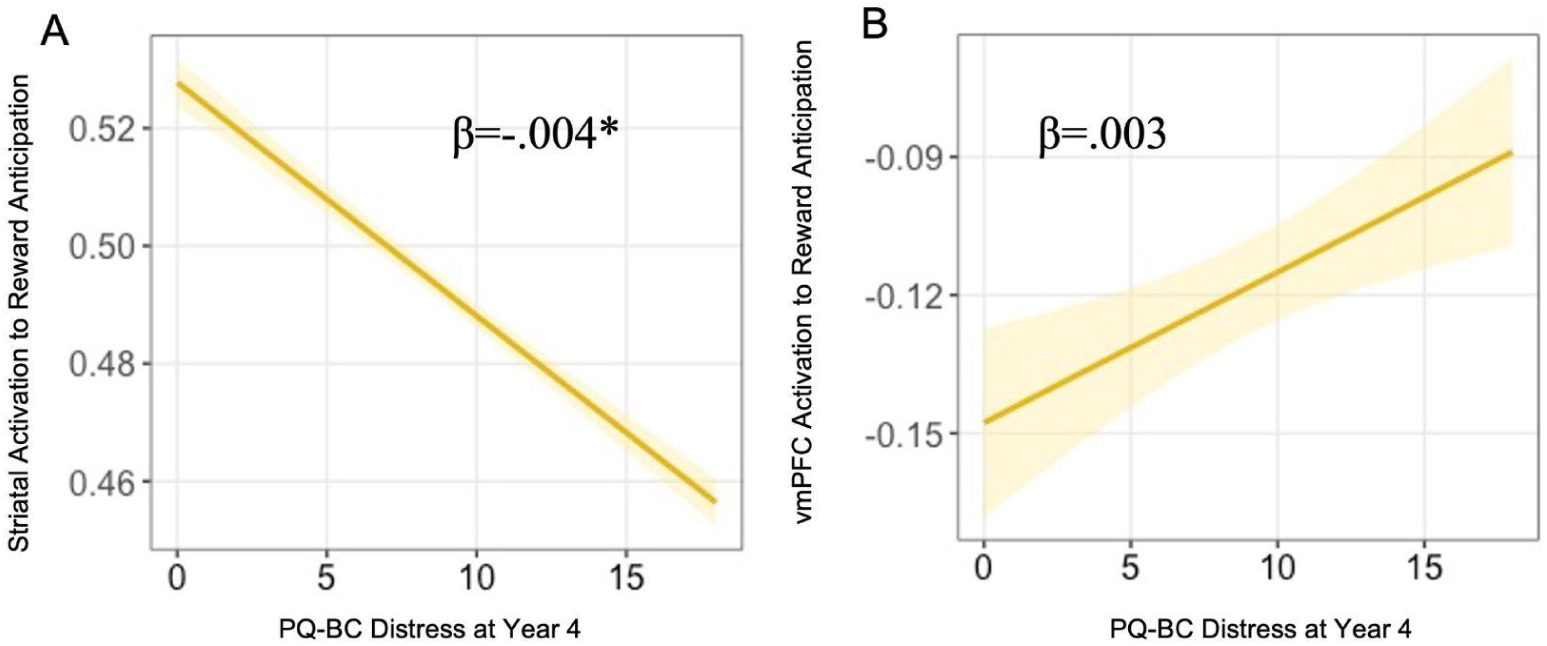
Results of models testing prospective associations between baseline activation during reward anticipation in A) the striatum and B) the ventromedial prefrontal cortex on the Monetary Incentive Delay Task across groups and psychotic-like experiences at year 4. Striatal activation was inversely associated with psychotic-like experiences at year 4 (A), whereas there was no significant association between vmPFC activation to reward anticipation at baseline and psychotic-like experiences at year 4 (B). Shading around the best-fit lines indicates standard error. Covariates: age, biological sex, parental education, income, mean framewise displacement, and baseline psychotic-like experiences as fixed effects with research and family unit (nested within site to account for twins and triplets in this study sample) as random effects. Abbreviations: PQ-BC=Prodromal Questionnaire - Brief Child Version; vmPFC=ventromedial prefrontal cortex. Significance: *p<.05, FDR corrected.

### MID Task Behavioral Performance

Models testing the association between psychotic-like experiences and behavioral performance on the MID task revealed reaction time to reward-predictive cues was positively associated with psychotic-like experiences in both groups (Figure 3 and Supplementary Table 4).

**Figure 3:**
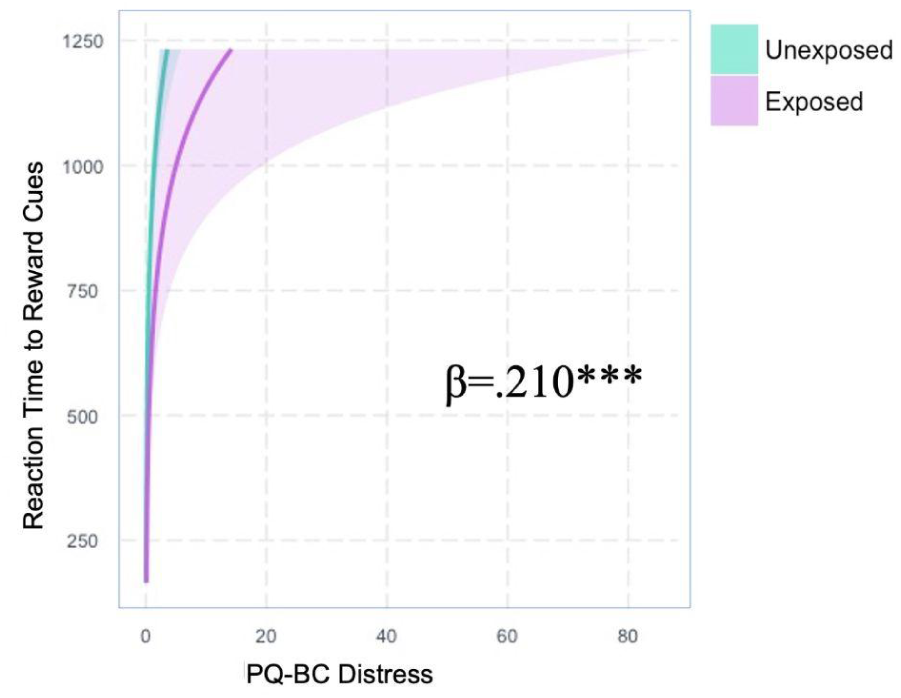
Associations between psychotic-like experiences and behavioral differences in reaction time during reward anticipation in the Monetary Incentive Delay Task by group status (exposed to cannabis prenatally versus unexposed) to high-magnitude reward-predictive cues. Reaction time was positively associated with psychotic-like experiences in both groups. Covariates: age, biological sex, parental education, and income as fixed effects, with research site and family unit (nested within site to account for twins and triplets in this study sample) as random effects. Participant ID was also included as a random effect to account for within-subject effects in longitudinal models. Beta values correspond to the relationship between psychotic-like experiences and behavioral measures (across groups), with stars indicating the strength of the relationship. Shading around the best-fit lines indicates standard error. Abbreviations: PQ-BC=Prodromal Questionnaire - Brief Child Version. Significance: ***p<.001, FDR corrected.

### Trait Measures

Psychotic-like experiences in PCE youth were associated with greater reward responsiveness, drive, excitement about reward receipt, negative urgency, and all four domains of impulsivity compared to unexposed youth, across time points (Supplementary Table 5 and Supplementary Figure 3). PCE youth scored higher across all trait-level measures. There were significant group-by-trait interactions for drive, negative urgency, lack of planning, and sensation seeking, such that relationships to psychotic-like experiences were stronger for PCE youth.

## 4. Discussion

This study is the first to examine longitudinal relationships between prenatal cannabis exposure (PCE), psychotic-like experiences, and behavioral and neural indices of reward anticipation over time in youth. We found, first, that previously observed associations between PCE and psychotic-like experiences in childhood persist through middle adolescence. Second, this study offers novel evidence implicating the brain’s reward pathways in the link between PCE and early signs of psychosis. Specifically: (1) Blunted neural response to reward anticipation is associated with psychotic-like experiences, with stronger effects observed in PCE youth; (2) Striatal hypoactivation at baseline predicts psychotic-like experiences in middle adolescence; (3) Dampened behavioral reward sensitivity is associated with psychotic-like experiences over time; and (4) Psychotic-like experiences are positively associated with trait-level measures of reward motivation and impulsivity across multiple timepoints, with stronger effects for PCE youth.

Our study extends recent work demonstrating an association between PCE and psychotic-like experiences in 9 and 10-year-olds exposed to cannabis after parental knowledge of pregnancy, which remains robust through middle adolescence after rigorously controlling for covariates.^8^ Not only did the association persist through middle adolescence, but the longitudinal association held regardless of whether a child was exposed before or after parental knowledge of pregnancy, indicating that even early exposure contributes to psychotic-like symptomatology later in childhood. The CB1 receptor is not expressed in the fetus until 5-6 weeks’ gestation, which corresponds approximately with knowledge of pregnancy;^33–35^ our findings support hypotheses suggesting there may still be indirect associations through endocannabinoid receptor expression in the placenta.^36^

In addition, we provide the first evidence that hypoactivation in the striatum during anticipation of reward at ages 9 to 10 is associated with psychotic-like experiences, with stronger effects for PCE youth; this baseline hypoactivation also predicts psychotic-like experiences in middle adolescence. Youth exposed to cannabis prenatally additionally showed more blunted activation in the vmPFC during reward anticipation compared to unexposed youth. Preclinical research shows adolescent and adult animals prenatally exposed to THC and other CB1 receptor agonists exhibit alterations in reward-related neural circuitry.^7^ Our results align with previous work demonstrating hypoactivation of the striatum during reward anticipation in adult patients with psychosis,^38^ extending this work to a youth sample with varying degrees of subthreshold psychotic symptoms. Additionally, we extend a previous study reporting blunted activation of the striatum to monetary reward anticipation in adult cannabis users,^20^ which may imply diminished orientation to a diminished future reward. The prefrontal cortex and value-based decision-making functions are not yet fully developed in this age group, which may explain the specificity of the association between psychotic-like experiences and blunted striatal activation, versus no association with vmPFC activation.^39–43^ That there were no associations between psychotic-like experiences and amygdala or insular activation implies specificity of these effects to the mesocorticolimbic pathway.^44^ These findings also extend recent work showing PCE is longitudinally associated with greater striatal resting state functional connectivity in children ages 9-12,^16^ in line with evidence of altered connectivity between the cortex and striatum during acute cannabinoid administration.^37^ Future work should elucidate whether the changing strength of frontostriatal connectivity during neurodevelopment is associated with youth psychopathology.

The endocannabinoid system, specifically the cannabinoid type 1 receptor (CB1), is the site of action of delta-9-tetrahydrocannabinol (THC), the primary psychoactive component of cannabis. CB1 receptors are preferentially expressed in key reward-related brain areas and regulate dopaminergic signaling.^45–47^ While a causal mechanism linking PCE to later onset of psychotic-like experiences has not yet been established, early alterations to the endocannabinoid-mediated dopaminergic system may contribute to vulnerability to psychosis in PCE youth.^48–53^ Our finding of prospective associations between baseline activation in reward-related brain regions and increased psychotic-like experiences in adolescence supports one potential mechanism through which PCE may lead to enhanced vulnerability to psychosis and produce long-term alterations in neural circuits involved in reward processing implicated in both acute psychotic-like symptoms and increased risk for the development of overt psychotic disorder. Moreover, blunted striatal activation may represent a biomarker of disrupted reward processing associated with subsequent psychotic-like experiences, observable as early as ages 9 and 10. While rates of substance use are low at the early ABCD study time points, future studies may wish to investigate whether this blunted striatal activation in childhood is also associated with later risk for substance use in adolescence.

The observed association between slower reaction times to reward-predictive cues and psychotic-like experiences in the present study may suggest dampened sensitivity to reward-related cues. Compared to typically developing youth, youth at clinical high risk for psychosis (CHR) display impaired behavioral performance during reward and cognitive tasks, associated with altered activation in reward-related brain regions and greater psychosis symptom severity, regardless of cannabis use history.^54–56^ Early THC exposure in both rodents and humans results in behavioral changes relevant to reward processing during adulthood such as increased risky decision-making and impulsivity, and an enhancement of the rewarding properties of drugs of abuse in rodents as measured by drug self-administration.^57–61^ Traits including increased reward responsiveness and impulsivity have previously been linked to adult cannabis use, although no causal mechanism has been established.^62^ In a rat model, high-dose THC exposure in adolescence results in risky decision-making and impulsivity in adulthood, as well as changes in reward learning, similar to findings seen in people who use cannabis.^63^

### Strengths and Limitations

This study leverages the largest prospective neuroimaging and behavioral study of adolescence to date to examine a key environmental risk variable associated with increased risk for psychosis. As the ABCD Study is an ongoing 10-year study, follow-up waves can be used to investigate whether childhood and adolescent psychotic-like experiences and reward processing deficits predict the persistence of psychopathology into later adolescence and young adulthood. Task-based functional imaging may offer a valuable biomarker for understanding mechanisms by which early cannabis exposure may contribute to later mental health outcomes.

This study also has certain limitations that should be noted. Cannabis use during pregnancy occurred roughly a decade earlier than these retrospective reports, and self-reported cannabis use was reliant on recall. Further, there is a lack of detailed information on cannabis potency, frequency of use, gestational age of exposure, and quantity of cannabis exposure during pregnancy in the ABCD study. While we rigorously controlled for multiple demographic and other covariates, unmeasured confounders may have a role. Direct studies of dose, frequency, and age-of-exposure are crucial for a better understanding of cannabis’ impact on the developing brain.

## 5. Conclusions

The results of this study demonstrate the significance of neurobehavioral markers of reward anticipation in the association between PCE and youth psychotic-like experiences. Ultimately, we provide important new evidence for lasting effects of early cannabis exposure on reward-related neural circuitry that may be a pathway impacting early psychosis risk.

## Data Availability

The study used openly available human data, originally located within the NIMH Data Archive ABCD Collection Release 5.1 (doi: 10.15154/z563-zd24)

