## Supplementary Materials for "Altered neurobehavioral reward response predicts psychotic-like experiences in youth exposed to cannabis prenatally"

**Table of Contents**

**Supplementary Methods and Results**

1. fMRI Data Acquisition and Analysis
2. Details on the Prodromal Questionnaire - Brief Child Version (PQ-BC)
3. Details on Prenatal Cannabis Exposure Measure
4. Details on the Monetary Incentive Delay Task
5. A priori Regions of Interest (ROIs)
6. Details on main linear mixed-effects models
7. Control Analysis: Amygdala and Insula
8. Details on Trait-Level Measures
   a. Behavioral inhibition system and behavioral activation system (BIS/BAS) scale.
   b. Urgency Premeditation Perseverance Sensation Seeking Positive Urgency Impulsive Behavior Scale (UPPS).
   c. Monetary Incentive Delay (MID) Post-Task Questionnaire

**Supplementary Figures**

Supplementary Figure 1: Schematic representation of data used across visits

Supplementary Figure 2: FSL parcellation of the striatum and vmPFC

Supplementary Figure 3: Models Examining Psychotic-like Experiences by Exposure Status in Trait-level Questionnaire Data.

**Supplementary Tables**

Supplementary Table 1: Summary of Models Examining Psychotic-like Experiences by Exposure Status in Trait-level Questionnaire Data.

Supplementary Table 2: Longitudinal Associations with Prenatal Cannabis Exposure

Supplementary Table 3: Rates of Participant Substance Use by Study Time Point.

Supplementary Table 4: Group Differences in the Baseline Association between Brain Activation in the Monetary Incentive Delay Task and Psychotic-like Experiences

Supplementary Table 5: Summary of Models Examining Group Differences in the Association between Behavioral Responses in the MID Task

**Supplementary Methods and Results**

1. **fMRI Data Acquisition and Analysis.** All participants were scanned on 3T scanners including Prisma (Siemens), Discovery MR750 (GE Healthcare), and Achieva dStream or Ingenia CX (Philips) with a 32-channel head coil. Adolescent Brain and Cognitive Development (ABCD) task-based functional assessment of the brain consists of three tasks assessed at baseline, 2-year follow-up, and four-year follow-up. This included the Monetary Incentive Delay (MID) task^1,2^, the primary task utilized in these analyses to probe behavioral and neural reward responsivity and sensitivity to reward-predictive cues^3^ in youth exposed to cannabis prenatally versus unexposed and associations to psychotic-like experiences in youth. The MID task used in the ABCD study measures the original CRAN ABCD RFA domains of reward processing, including anticipation and receipt of reward and losses, and trial-by-trial motivation in speeded responses to win or avoid loss. Functional magnetic resonance imaging (fMRI) preprocessing was conducted by ABCD’s Informatics Center.^4^
2. **Details on the Prodromal Questionnaire - Brief Child Version (PQ-BC).** Participants were asked to indicate whether they experienced each item (yes/no). If they endorsed a given psychotic-like experience (PLE), they were further asked whether the experience bothered them (yes/no). If the experience was bothersome, participants then rated the level of distress on a 5-point Likert scale. A summary score for PQ-BC Distress was calculated by adding the total number of endorsed items weighted by distress.^5–9^ Specifically, each item was scored as follows: 0 for not experiencing the PLE, 1 for experiencing the PLE with no distress, and 2-6 for distressing psychotic-like experiences (PLEs), with the distress score added to the item score. The PQ-BC Distress score was utilized as the measure of PLEs in all analyses.
3. **Details on Prenatal Cannabis Exposure Measure.** ABCD study prevalence estimates of self-reported prenatal cannabis use align with toxicology-based prevalence estimates from national data sets collected during the years these children were born, supporting accuracy of self-report.^10^
4. **Details on the Monetary Incentive Delay Task.** For our analyses, data from participants with information on PCE and passed functional neuroimaging MID task quality control recommendations were included (see <https://wiki.abcdstudy.org/release-notes/imaging/quality-control.html>. The Monetary Incentive Delay (MID) task is a well-validated fMRI probe designed to measure domains of reward processing ^1,3^. It reliably activates reward-related brain circuitry, is sensitive to developmental^10–12^ and addiction-related effects (i.e., over-activation in reward-related regions in individuals with substance use disorders),^13–16^ and has strong within-subject test-retest reliability over time.^15^ The ABCD Data Analysis, Informatics and Resource Center conducts centralized processing of MRI data across ABCD sites, using the Multi-Modal Processing Stream.^4^ Analyses utilized summary metrics of task-based activation and behavioral responses available in the tabulated data. Due to our a priori hypotheses, reward-related brain regions - specifically, the striatum and ventromedial prefrontal cortex (vmPFC) - were tested for associations, with amygdala and insula tested in secondary analyses. We used the ABCD’s inclusion recommendations for each participant, based on the quality of their task-based imaging scans and included participants if they passed these inclusion criteria (n=11,368 at baseline; 2-year follow-up n=7,928, and 4-year follow-up n=2,982). A detailed description of inclusion criteria are discussed in NDA 4.0 MRI Quality Control Recommended Inclusion (http://dx.doi.org/10.15154/1523041); in brief, these criteria take into account factors including imaging quality control and task performance. Contrast-specific exclusion thresholds for beta values in a single ROI were calculated as the mean ± 3*standard deviations of beta values within the sample for that contrast-ROI. The participant-events marked for potential exclusion had 25% or more of their ROIs detected as outliers.

Lasting approximately 12 minutes, the MID task involves two runs, each containing 50 trials (10 per trial type).^3^ Participants see incentive cues indicating whether they can win big (+$5; large reward condition), lose large (-5$; large loss condition), win small (-$.20; small reward condition), or lose small (-$.20, small loss condition). After an anticipatory period, a target appears on the screen and participants are required to make a button press as quickly as they can. Participants are then shown feedback indicating their subsequent reward or loss. Monetary reward (on average $21) is distributed to participants at the end of each task visit. Data were analyzed using the contrast for large reward cue anticipation versus neutral cue anticipation trials (large reward). Each participant is compensated for their specific performance and earns at least $1 to ensure motivation during their scans.^3^ The contrast of the large magnitude reward (+$5) versus the neutral cue was used as the measure of neural activation to reward anticipation in neuroimaging analyses, in line with previous studies demonstrating high-magnitude reward condition elicits the most reliable reward response.^1,11–13^ Response time to high-magnitude reward cues was used in behavioral analyses as the measure of reaction time.^14^

5. **A priori Regions of Interest (ROIs)**: We assessed neural activation in reward-related brain regions, specifically activation in the striatum and ventromedial prefrontal cortex (vmPFC), during reward anticipation using regions of interest. These regions are commonly implicated in reward processing, as well as in psychosis and CUD, and reliably activated by the MID task.^2,11,15^ Striatal hypoactivation during the anticipation phase of the MID task has previously been shown in both adults with CUD^16^ and adults with schizophrenia.^17^
6. **Details on main linear mixed effects models.** The PQ-BC data are zero-inflated and positively skewed; therefore, models with PLEs as the dependent variable utilized Poisson distribution general linear models (log link function). Longitudinal models tested across the three waves of data: baseline, 2-year follow-up visit, and 4-year follow-up visit. To test for differences regarding whether a child was exposed to cannabis before or after parental knowledge of pregnancy,^18^ we re-ran linear mixed models with PLEs as the dependent variable and cannabis exposure group models treating the independent variable as a three-level factor (exposed after parental knowledge of pregnancy versus exposed before parental knowledge of pregnancy versus unexposed). Based on findings from this initial analysis, we collapsed across exposure groups for all subsequent analyses.
7. **Control Analysis: Amygdala and Insula.** As a control analysis to test the specificity of results to the dopaminergic mesocorticolimbic pathway, we also re-ran these models with amygdala and insula ROIs; though not primary elements of this pathway, both regions are involved in the mechanisms of addiction, substance use, and reward-related decision making.^19–21^ For example, the insula plays a key role in the representation of interoceptive effects of drugs and their integration with attention, executive function, and emotion,^22^ and the amygdala contributes to the pathogenesis of addictive disorders and drug-seeking and self-administrative behaviors.^23,24^ We hypothesized that activation in the amygdala and insula would not be associated with PLEs.
8. **Description of Self-Report Trait-level Measures.** Self-report trait measures included reward responsiveness and drive, as measured by the behavioral inhibition system and behavioral activation system (BIS/BAS) scale^25,26^; excitement about rewards, as measured by a post-MID task questionnaire; and four measures of impulsivity, including: 1) negative urgency, 2) lack of planning, 3) sensation seeking, and 4) positive urgency as measured by the Urgency Premeditation Perseverance Sensation Seeking Positive Urgency Impulsive Behavior Scale (UPPS).^27^ These self-report measures were collected at baseline, 2-year follow-up, and 4-year follow-up; all three time points were included in our analyses.
   a. **Behavioral inhibition system and behavioral activation system (BIS/BAS) scale**. Reward Responsiveness and Drive measures from the Youth Behavioral Inhibition/Behavioral Approach System Scales (BIS/BAS), a version of Carer and White’s (1994) BIS/BAS Scale adapted for youth,^28^ were used in trait-level analyses of reward responsiveness and drive. Reward Responsiveness includes items such as, “When good things happen to me, it affects me strongly.” Drive includes items like “I go out of my way to get things I want”.
   b. **Urgency Premeditation Perseverance Sensation Seeking Positive Urgency Impulsive Behavior Scale (UPPS).** The UPPS impulsivity scale^29^ is a 46-item inventory created to measure four distinct personality pathways to impulsive behavior. The inventory was derived through a factor-analytic method that included well-known impulsivity scales. The youth version of the UPPS used in the ABCD study demonstrates the same five-factor structure as the adult version and shows good convergent and discriminant validity with relevant personality, psychopathology, and neurocognitive measures.^27^ Negative urgency is defined by the tendency to act without thinking. Positive urgency is the tendency to act rashly under extreme positive emotions. Sensation-seeking is the tendency to seek out novel and thrilling experiences. Lack of planning is the tendency to act without thinking.
   c. **Monetary Incentive Delay (MID) Post-Task Questionnaire.**^3^ Participants were surveyed following the MID task regarding their experience during the task. We used post-MID task measures of excitement about rewards from this survey. Specifically, we used the item which asked “how excited were you when you saw a circle saying you could win $5?” to measure excitement about rewards. Answers were rated on a 5-point Likert scale (1 = Very slightly or not at all [excited]; 2 = A little; 3 = Moderately; 4 = Quite a bit; 5 = Extremely).

**Supplementary Table 1:** Rates of Participant Substance Use by Study Time Point.

| Substance Use (mean (SD)) | Prenatal Cannabis Exposure | No Prenatal Exposure |
| --- | --- | --- |
| Two-Year Follow-up Visit |  |  |
| Cannabis Use (last year) | 14.50 (4.04) | 18.73 (46.77) |
| Tobacco Use (last year) | 3.40 (4.55) | 3.48 (5.69) |
| Alcohol Use (last month) | 1.75 (.89) | 1.63 (2.02) |
| Four-Year Follow-up Visit |  |  |
| Cannabis Use (last year) | 29.76 (76.89) | 14.56 (66.09) |
| Tobacco Use (last year) | 13.97 (65.48) | 12.35 (42.65) |
| Alcohol Use (last month) | 2.92 (9.56) | 3.72 (11.87) |

We analyzed self-report and clinical interview data from the ABCD Study. Youth (2-year follow-up n=214, M_age_ =10.20±7.12; 4-year follow-up n=7,842, M_age_=12.96±.63) completed a timeline follow-back substance use assessment.^4,30–32^ Youth were interviewed about their use of illicit substances in both the last 30 days and last year. Alcohol is measured as combined count of total standard units of alcohol during the past month, whereas nicotine (including but not limited to cigarettes, ENDS, smokeless tobacco, cigars, hookah, pipe, nic replacement) and cannabis use (including but not limited to smoked/vaped cannabis flower, blunts, edibles, smoked/vaped concentrates, cannabis alcohol drinks, cannabis tinctures, synthetic cannabis) is measured as the total days of any type of use in the last year. For these analyses, alcohol, nicotine, and cannabis use were cumulatively scored and are reported as means and standard deviations by exposure status. Two- and four-year follow-up visit use rates are reported, owing to limited endorsement of ever having tried illicit substances at the baseline time point among ABCD youth participants.

**Supplementary Table 2:** Longitudinal associations of psychotic like experiences (PLEs) with prenatal cannabis exposure by two- (youth exposed to cannabis prenatally versus unexposed youth) and three-level factor [exposed after parental knowledge of pregnancy (PCE-A) versus exposed before parental knowledge of pregnancy (PCE-B) versus unexposed (UE)]

|  | **Exposed vs. Unexposed (across exposure status groups)** | | **PCE-A vs. UE** | | **PCE-B vs. UE** | | **PCE-B vs. PCE-A** | |
| --- | --- | --- | --- | --- | --- | --- | --- | --- |
|  | std. β | std. CI | std. β | std. CI | std. β | std. CI | std. β | std. CI |
| **Psychotic-like Experiences** | .370*** | [0.21, 0.52] | .470*** | [0.22, 0.73] | .260** | [0.07, 0.45] | -.014 | [-0.45, 0.17] |

Prenatal cannabis exposure is significantly associated with psychotic-like experiences, across all timepoints. There were no statistically significant differences in PLEs between those exposed to cannabis before and after parental knowledge of pregnancy.

**Covariates:** age, biological sex, parental education, income, birth weight, prenatal exposure to tobacco or alcohol before or after maternal knowledge of pregnancy, unplanned pregnancy, and prenatal vitamin use, with research and family unit (nested within site to account for twins and triplets in this study sample) as random effects.

**Reference categories:** Exposed; PCE-A; PCE-B; PCE-B. **Abbreviations:** UE=Unexposed; PCE-A=exposed to cannabis after parental knowledge of pregnancy; PCE-B=exposed to cannabis before parental knowledge of pregnancy only. **Significance***:* *q<.05, **q<.01, ***q<.001, FDR corrected

**Supplementary Table 3:** Group Differences in the Baseline Association between Neural Activity in the Monetary Incentive Delay Task and Psychotic-like Experiences

|  | **Effect of Psychotic-Like Experiences** | | **Effect of Group (Exposed vs. Unexposed)** | | **Group * Region Interaction** | |
| --- | --- | --- | --- | --- | --- | --- |
|  | std. β | std. CI | std. β | std. CI | std. β | std. CI |
| **Large Reward Condition** | | | | | | |
| **Striatum** | **-.050*** | [-0.10, -0.01] | **.060**** | [0.02, 0.11] | -.005 | [-0.14, 0.21] |
| **vmPFC** | -.020 | [-0.07, 0.02] | **.070**** | [0.03, 0.11] | .004 | [-0.17, 0.17] |

Striatal activation to reward-predictive cues during the MID Task is inversely associated with reported psychotic-like experiences. However, there is no significant association between vmPFC activation and psychotic-like experiences. There are significant main effects of group; PCE youth showed more blunted activation in both the striatum and vmPFC during reward anticipation compared to unexposed youth. There are no significant group-by-region interactions. **Reference level for group effect:** youth exposed to cannabis prenatally. **Covariates:** age, biological sex, parental education, income, and mean framewise displacement as fixed effects, with research and family unit (nested within site to account for twins and triplets in this study sample) as random effects. **Significance:** *q<.05, **q<.01, ***q<.001, FDR corrected. **Abbreviations:** vmPFC=ventromedial prefrontal cortex.

**Supplementary Table 4:** Summary of Models Examining Group Differences in the Association between Behavioral Responses and Psychotic-like Experiences in the MID Task

|  | **Psychotic-Like Experiences** | | **Effect of Group (Exposed vs. Unexposed)** | | **Group * RT Interaction** | |
| --- | --- | --- | --- | --- | --- | --- |
|  | std. β | std. CI | std. β | std. CI | std. β | std. CI |
| **Large Reward Condition** | | | | | | |
| **RT (in ms)** | **.210***** | [0.18, 0.23] | 0.09 | [-0.06, 0.25] | .25 | [-0.08, 0.20] |

Associations between psychotic-like experiences and reaction time in the Monetary Incentive Delay Task by group status (exposed to cannabis prenatally versus unexposed) to high-magnitude reward-predictive cues. Reaction time was positively associated with psychotic-like experiences in both groups. **Abbreviations:** RT= Reaction time (in milliseconds). **Covariates:** age, biological sex, parental education, and income as fixed effects with research and family unit (nested within site to account for twins and triplets in this study sample) as random effects. **Significance:** *q<.05, **q<.01, ***q<.001, FDR corrected.

**Supplementary Table 5:** Summary of models examining associations between self-reported reward-responsivity and impulsivity-related traits and psychotic-like experiences over time, by prenatal cannabis exposure status.

|  | **Main Effect of Psychotic-Like Experiences (Regardless of Exposure Status)** | | **Effect of Group (Exposed vs. Unexposed)** | | **Group * Trait Interaction** | |
| --- | --- | --- | --- | --- | --- | --- |
|  | std. β | std. CI | std. β | std. CI | std. β | std. CI |
| Reward Responsiveness | **.25***** | [0.22, 0.27] | **.32**** | [0.17, 0.46] | .01 | [-0.07, 0.09] |
| Drive | **.22***** | [0.20, 0.24] | **.35***** | [0.21, 0.50] | **-.14***** | [-0.22, -0.06] |
| Excitement about Rewards | **.10***** | [0.08, 0.13] | **.32*** | [0.17, 0.46] | .01 | [-0.07, 0.10] |
| Negative Urgency | **.40***** | [0.38, 0.42] | **.41***** | [0.38, 0.42] | **-.14**** | [-0.22, -0.06] |
| Lack of Planning | **.12***** | [-0.22, -0.06] | **.43***** | [0.28, 0.57] | **-.07***** | [-0.15, 0.01] |
| Sensation Seeking | **.15***** | [0.13, 0.18] | **.35***** | [0.20, 0.49] | **-.11**** | [-0.20, -0.03] |
| Positive Urgency | **.31***** | [0.29, 0.34] | **.33***** | [0.19, 0.47] | -.05 | [-0.13, 0.02] |

Psychotic-like experiences are associated with trait-level measures of reward responsivity and motivation, with significant main effects of group for all measures (**Reference level for group effect**: exposed to cannabis prenatally). There are significant interactions for drive, negative urgency, lack of planning, and sensation seeking such that effects on PLEs are stronger for youth exposed to cannabis prenatally. Reward responsiveness and drive were measured using the Behavioral inhibition system and behavioral activation system (BIS/BAS) scale; excitement about potential task rewards was measured using the MID post-task questionnaire; negative urgency, lack of planning, sensation seeking, and positive urgency were measured using the Urgency Premeditation Perseverance Sensation Seeking Positive Urgency Impulsive Behavior Scale (UPPS). This includes data from baseline (n=11,845), 2-year follow-up visit (n=10,941), and 4-year follow-up visit (n=4,735). See Supplementary Methods and Results, section 4 for detailed information on these measures. **Abbreviations:** UE=Unexposed. **Covariates:** age, biological sex, parental education, and income as fixed effects with research site and family unit (nested within site to account for twins and triplets in this study sample) as random effects. Participant ID was also included as a random effect to account for within-subject effects in longitudinal models. **Significance:** *q<.05, **q<.01, ***q<.001, FDR corrected.

**Supplementary Figure 1:** Schematic representation of data used across visits.

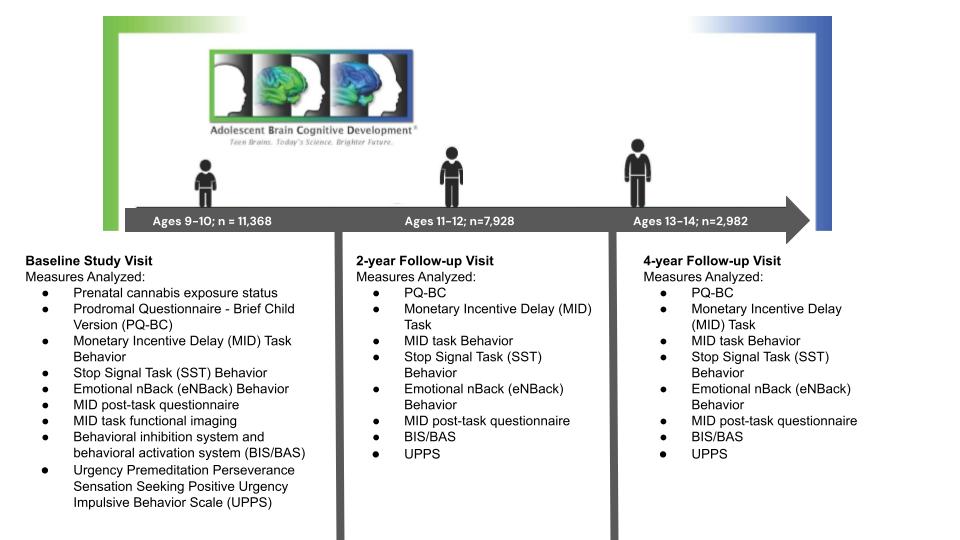

Study schematic depicting the three time points of the Adolescent Brain and Cognitive Development (ABCD) Study utilized in these analyses. This schematic outlines the longitudinal design and the progression of measures across the study to identify early behavioral and biomarkers of the onset of psychotic-like experiences following prenatal cannabis exposure. The sample size listed at each study time point reflects data analyzed for this study.

**Supplementary Figure 2:** Freesurfer parcellation of the striatum (blue) and ventromedial prefrontal cortex (red).

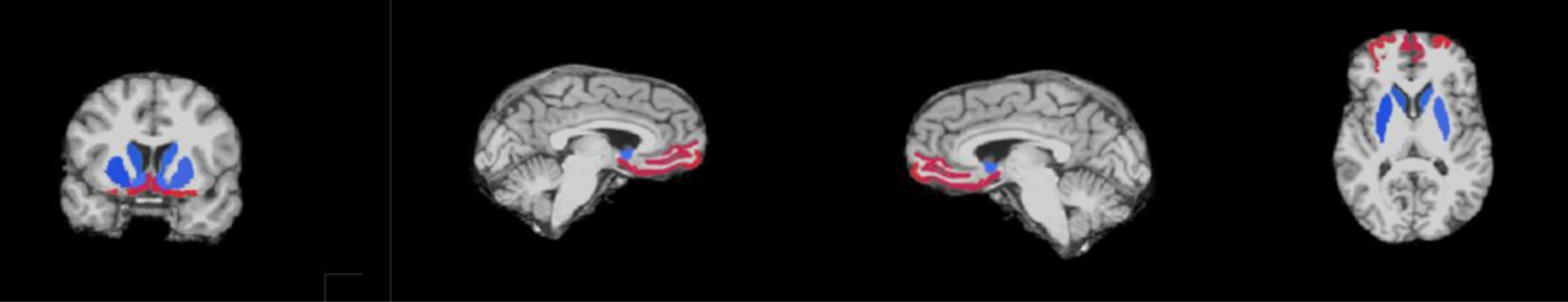

Brain regions of interest (ROIs) derived from FreeSurfer. Cortical surface reconstruction and subcortical segmentation were performed using FreeSurfer v5.3, which includes tools for estimation of various measures of brain morphometry and uses routinely acquired T1w MRI volumes.^33–38,38^ The FreeSurfer package has been validated for use in children^39^ and used successfully in large pediatric studies.^40^ The striatum is marked in blue, and the ventromedial prefrontal cortex is marked in red. These regions are critical in the study of early biomarkers of psychotic-like experiences following prenatal cannabis exposure. CB1 receptors are distributed and preferentially expressed in key reward-related brain areas including the striatum and ventromedial prefrontal cortex (vmPFC) and regulate dopaminergic signaling.^41–43^ The MID task is a robust activator of the striatum and vmPFC, demonstrating validity as a probe of reward responding.^2,3,11^

**Supplementary Figure 3:** Models examining associations between self-reported reward-responsivity related traits and psychotic-like experiences, by prenatal cannabis exposure status.

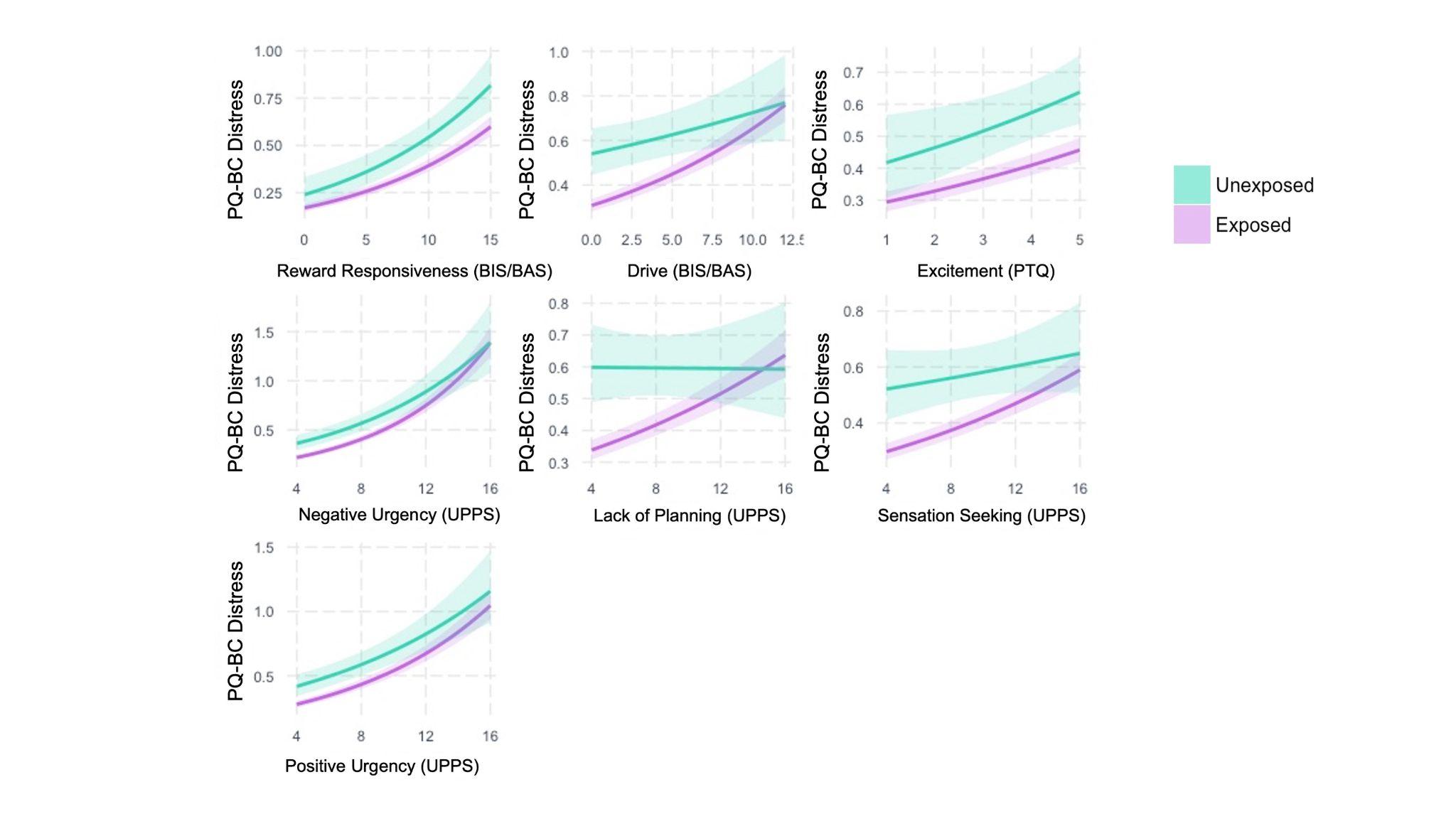

Psychotic-like experiences are associated with trait-level measures of reward responsivity and motivation across all time points, with significant main effects of group (exposed to cannabis prenatally versus unexposed). There are significant interactions for drive, negative urgency, lack of planning, and sensation seeking, such that effects are stronger for youth exposed to cannabis prenatally. Reward responsiveness and drive were measured using the Behavioral inhibition system and behavioral activation system (BIS/BAS) scale; excitement about potential task rewards was measured using the MID post-task questionnaire;^3^ negative urgency, lack of planning, sensation seeking, and positive urgency were measured using the Urgency Premeditation Perseverance Sensation Seeking Positive Urgency Impulsive Behavior Scale (UPPS). This includes data from baseline (n=11,845), 2-year follow-up visit (n=10,941), and 4-year follow-up visit (n=4,735). See Supplementary Methods and Results, section 4 for detailed information on these measures. Shading around the best-fit lines indicates standard error. **Abbreviations:** PQ-BC=Prodromal Questionnaire - Brief Child Version; PTQ=Post-task questionnaire (from the Monetary Incentive Delay task). **Covariates:** age, biological sex, parental education, and income as fixed effects, with research site and family unit (nested within site to account for twins and triplets in this study sample) as random effects. Participant ID was also included as a random effect to account for within-subject effects in longitudinal models.
